# The Missing Science: Epidemiological data gaps for COVID-19 policy in the United States

**DOI:** 10.1101/2021.02.11.21251602

**Authors:** Rajiv Bhatia, Isabella Sledge, Stefan Baral

## Abstract

We report on a scoping study of COVID-19 epidemiological data available for COVID-19 policy and management decisions for U.S. settings. We synthesize current US Centers for Disease and Control and Prevention (CDC) estimates for parameter of infectious transmission, infection severity, and disease burden, and summarize epidemiologic contributions to these parameters published by CDC-affiliated investigators through Oct 30, 2020. Authoritative estimates of most infectious transmission and infection severity parameters exist but rely primarily on data from studies conducted in non-U.S. populations. Estimates of secondary infection risks for household, workplace, school, or other community settings are lacking and estimates of the clinical fraction remain uncertain. The CDC reports multiple disease incidence and prevalence measures at national and state geographies, including some measures disaggregated by age group, race/ethnicity and sex; however, nationally uniform disease burden measures are not available at the sub-state level or for sub-populations defined by exposure setting, limiting opportunities for targeted interventions. CDC-affiliated investigators authored 133 quantitative studies on COVID-19 through Oct 30, 2020; however only 34 employed analytic methods. The remainder were descriptive. Of the 34 analytic studies, eleven reported on risk factors for infection, seven reported on risk factors for severe disease, three on symptomatic infections, three reported secondary infection risks, and four reported on indirect pandemic effects. Gaps remain in the epidemiological data required for an efficient and equitable public health policy response to COVID-19. The existence of these gaps one year after the onset of the COVID-19 pandemic underscores the need for standardizing data collection and research priorities and protocols in the context of a rapidly emerging infectious disease epidemics.

## Introduction

In any emerging infectious disease epidemic, efficient and defensible disease control interventions will require timely and complete epidemiological data. The scope and severity of the US COVID-19 epidemic and its enormous social costs makes the need for actionable epidemiologic data particularly acute. Yet, one year after the discovery of the SARS-CoV-2 virus, the US Centers for Disease Control (CDC) acknowledges that “information about the biological aspects of SARS-CoV-2 and epidemiological characteristics of COVID-19 remain limited….” [1] Insufficient data on transmission mechanisms, setting and activity specific risks, and population-specific disease burden may have reduced the effectiveness, efficiency, and equity of the public health response. [2]

An established literature on emerging respiratory virus epidemics provides a framework for evaluating the adequacy of Covid-19 data required for policy and management decisions. Lessons learned from prior epidemics including H1N1 influenza (H1N1), Severe Adult Respiratory Syndrome (SARS-CoV-1), and Middle-East Respiratory Syndrome (MERS) all inform CDC pandemic plans for emerging influenza viruses. The CDC’s pandemic interval framework [3] and the pandemic severity assessment framework [4] explicitly spell out the scientific and data needs for assessing the risks posed by emerging respiratory viral infections. Reviews critically examining the epidemiology of the H1N1 pandemic response elaborate on these data demands, [5,6,7,8] which were re-iterated at the onset of the COVID-19 pandemic. [9,10]

A scoping study is a method to examine the extent, range, nature of evidence or research activities in a particular field and to identify research gaps. [11] Here, we report on a scoping study of COVID-19 epidemiologic parameters for infectious transmission, infection severity, and disease burden which, collectively, inform the threat level posed by an emerging respiratory virus epidemic and guide an efficient, proportional and equitable public health response. [8]. We first synthesize current authoritative federal government appraisals for each of these parameters. We then summarize the methods and outcomes of epidemiologic investigations published by CDC-affiliated investigators informing the aforementioned parameters. We focus specifically on CDC appraisals and investigations because of the agency’s unique access to the data required for COVID-19 epidemiology, its partnerships with state and local public health authorities, and its leadership role in pandemic management. We identify knowledge gaps and suggest opportunities to improve the evidence base for COVID-19 mitigation.

## METHODS

Table 1 summarizes the parameters for infectious transmission, disease severity, and disease burden that we sought along with their typical data sources.

**Table 1.** Epidemiological data required for emerging respiratory virus epidemic management

| Measure | Definition | Information value | Typical source |
| --- | --- | --- | --- |
| **Infectious Transmission** |  |  |  |
| Basic reproductive number | The expected number of secondary cases directly generated by one case | Potential speed of epidemic growth | Calculated from contact rate, secondary infection risk, and infectious period or from the growth rate of the early disease incidence curve |
| Growth rate | Change per unit time (acceleration or deceleration) of the incidence rate | Current trajectory of epidemic growth | Population disease monitoring |
| Fraction of the population susceptible | Proportion of the population who do not have immunity to the infection or to disease | Targeting control measures | Studies of immunity such as seroprevalence of antibodies |
| Incubation period | Interval between infection and the development of symptoms | Timeframe for prevention of secondary infection | Transmission studies |
| Duration of infectiousness | Viral load and duration in symptomatic and asymptomatic people | Timeframe for prevention of secondary infection | Transmission studies |
| Serial interval | Interval between development of symptoms in a case and an infected contact | Necessary for capturing the $R_0$ | Transmission studies |
| Fraction of pre-symptomatic transmission | Proportion of infections spread by persons who appear well but are infected and later develop symptoms | Timeframe for prevention of secondary infection | Transmission studies |
| Secondary infection risk (SIR or alternatively, secondary attack rate) | Proportion of exposed people who become ill particular setting (household, school workplace) | Targeting control measures | Transmission studies |
| **Infection Severity** |  |  |  |
| Clinical fraction | Proportion of infected people who become ill | Predicting number of transmitted infections that will result in illness; adopting a proportional response | Household or contact tracing transmission studies |
| Case or infection hospitalization ratios | How many people who become infected require hospitalization | Predicting the scale and scope of health care services; adopting a proportional response | Population surveillance<br>Cohort studies<br>Large transmission studies |
| Case or infection fatality ratio | How many infected people die | Adopting a proportional response | Reported hospital data. Death records |
| Severity risk factors | Risk factors, including ethnicity, income, comorbidities, healthcare access, social characteristics, environmental conditions, affecting vulnerability to disease | Targeting control measures | Case control studies<br>Syndromic surveillance |
| **Disease Burden** |  |  |  |
| Incidence rate | Number of new cases of illness, hospitalization or death in a population per unit time | Is the disease accelerating or slowing down and where | Syndromic surveillance<br>Serial prevalence studies<br>Disease hospitalization rates<br>Disease mortality rates |
| Point or period prevalence | Proportion of the population that is a current case a point or period in time | Current level of active infection and transmission | Symptom and test-based surveys |
| Community attack rate / cumulative incidence | Number of new cases of disease during specified time interval | Population disease burden & remaining population susceptible | Cohort studies, statistical estimation |

Transmission studies, which observe individuals with confirmed infection and the people they infect secondarily, provide data on the mechanism of transmission (e.g. person to person through respiratory secretions), the incubation period (how long after infection symptoms appear), the generation interval (the time between a person becoming infected and subsequently infecting someone else), the infectivity period (how long an infected person can spread the illness), and the risk of infection among exposed contacts in various settings (secondary infectious risks/secondary attack rates). Transmission studies also identify populations relatively more susceptible to infection as well as groups or settings relatively more likely to contribute to transmission. [12] Additionally, transmission studies can quantify pre-symptomatic transmission and the proportion of infections that are symptomatic (the clinical fraction). These latter two parameters will, in part, determine the feasibility of isolation and contact tracing strategies. [13,14] Optimal understanding of transmission and an equitable mitigation response requires that studies include diverse community settings (e.g., household, workplace, school, and congregate living such as long-term care facilities and shelters).

Infection severity measures include estimates of the proportion of infected individuals who require hospitalization or who die. Estimates of infection severity depend upon probable and confirmed case definitions and thus on the surveillance systems that ascertain infection and infection-related morbidity and death. Severity measures are vulnerable to many biases and may vary by place and over time as surveillance methods, case definitions, testing strategies, clinical understanding, and prevention efforts evolve. Cohort studies including large case ascertained studies (where households are enrolled after an index case is identified) with adequate follow up can provide reliable estimates of disease severity measures in addition to informing transmission risks. [8]

Disease burden is the product of infection transmission and severity contextualized by socio-demographic and environmental risk factors and their disparities. Measures of disease burden include the incidence and prevalence of illness, numbers of people hospitalized, and infection-related deaths. Systematically monitoring health care visits for infection-related illness is one reliable indicator of the disease burden if such surveillance is combined with routine testing of a representative sample of patients visits. [8] Optimally, data on the incidence of infection-related illness should be available in real time and disaggregated by population subgroup, setting, severity and patient characteristics to inform timely, targeted community mitigations and anticipation of healthcare needs.

The CDC’s Pandemic Severity Assessment Framework (PSAF), an element of the national pandemic strategy, integrates the aforementioned transmission, severity and disease burden parameters into a qualitative assessment of the threat posed by an emerging infection. [4] Such risk assessments should be updated as the epidemic evolves, and data improves. Table S1 (Supplemental Materials) illustrates CDC’s application of the PSAF for 2009 Influenza H1N1. [4]

We accessed public CDC webpages (See list in supplemental Materials, Table S2) to find current federal estimates of SARS-CoV-2 transmission, infection severity, and disease burden parameters and their data sources. We also searched specifically for documents containing applications of pandemic risk assessment tools, such as the PSAF.

To evaluate original contributions to COVID-19 epidemiology by CDC-affiliated investigators, we first reviewed all studies in the collection of COVID-19 research in the Morbidity and Mortality Weekly Report (MMWR)—a principal vehicle for publishing studies conducted by CDC investigators and their state partners—as well as the CDC COVID-19 Science update webpage. We further searched PubMed for studies with COVID-19 in the title or abstract, a medical subject heading (MeSH) for epidemiology, and CDC as an affiliation. Additionally, we examined the COVID-19 research collections in Emerging Infectious Disease, Clinical Infections Disease (CID), the Journal of the American Medical Association (JAMA), and the New England Journal of Medicine (NEJM). We included studies published from 1 Jan to 30 Oct 2020. We excluded studies that did not present quantitative data.

We classified each included study as either descriptive or analytic and then abstracted published information on study design, data source, setting, population, and outcomes. (Supplemental Materials, Table S3) We considered a study descriptive if it did not evaluate or formally test a hypothesis using a well-established epidemiological method. We classified studies that quantified risk of infection, disease severity, or transmission through an established epidemiological method as analytic. If a study reported both descriptive and analytic components, we classified it as analytic.

We subcategorized descriptive studies by their design as: case study, case series, cluster or outbreak investigation, cross-sectional or ecologic survey. We categorized the designs of analytic studies as either cross-sectional, case-control, ecologic prospective cohort, or retrospective cohort designs. We identified each study’s data source as either passive, sentinel, syndromic, or active surveillance programs, administrative program records, medical records, or survey instruments. We also categorized each study with regards to its setting and population.

We abstracted study outcomes as reported by the investigators. For each study other than those in the case studies and the case series, cluster and outbreak categories investigations, we identified outcomes contributing to any of the key aforementioned epidemiologic parameters.

## RESULTS

Table 2 synthesizes CDC’s current estimates of epidemiologic parameters for SARS-CoV-2 transmission, infection severity and disease along with the estimates referenced sources.

**Table 2.** CDC estimates for SARS-CoV-2 infection transmissibility, severity and disease burden measures

| Measures | Estimate (range) | Estimate date | CDC Webpage | Sources referenced by CDC | Referenced source study setting(s) |
| --- | --- | --- | --- | --- | --- |
| **Transmissibility** |  |  |  |  |  |
| Reproductive number | 2.5 (2.0 – 4.0) | 10 Sep 2020 | COVID-19 Pandemic Planning Scenarios [15] | Li et al. [16] | Multiple settings |
| Susceptibility | 100% | 10 Sep 2020 | COVID-19 Pandemic Planning Scenarios [15] | Not sourced |  |
| Mean incubation period | ~ 6 days (mean) | 10 Sep 2020 | COVID-19 Pandemic Planning Scenarios [15] | McAloon et al.[17] | China |
| Mean serial interval | ~ 6 days (mean) | 10 Sep 2020 | COVID-19 Pandemic Planning Scenarios [15] | He et al.[18] | China |
| Percentage of transmission occurring prior to symptom onset | 50% (30 - 70) | 10 Sep 2020 | COVID-19 Pandemic Planning Scenarios [15] | He et al.[18]; Casey et al.[19] | China, Northern Italy |
| Duration of infectiousness | < 10 days after symptom onset | 19 Oct 2020 | Duration of Isolation and Precautions for Adults with COVID-19 [20] |  |  |
| Secondary infection risk (SIR) | No estimates |  |  |  |  |
| **Infection Severity** |  |  |  |  |  |
| Percent of infections that are asymptomatic | 40% (10 – 70%) | 10 Sep 2020 | COVID-19 Pandemic Planning Scenarios [15] | Byambasuren et al. [21]; Poletti et al.[89] | Multiple settings |
| Infection hospitalization ratio | No estimates |  |  |  |  |
| Infection fatality ratio | 0-19 years: 0.00003;<br>20-49 years: 0.0002;<br>50-69 years: 0.005;<br>70+ years: 0.054 | 10 Sep 2020 | COVID-19 Pandemic Planning Scenarios [15] | Hauser et al.[22] | European Union |
| Hospital fatality ratio | 18-49 years: 2.4%; 50-64 years: 10.0%; ≥65 years: 26.6% | 10 Sep 2020 | COVID-19 Pandemic Planning Scenarios [15] | COVID-NET [23] | USA, 13 States |
| Rate ratio for hospitalization by age | 0-4 years: 4x lower<br>5-17 years: 9x lower<br>18-29: comparison<br>30-39 years: 2x higher<br>40-49 years: 3x higher<br>50-64 years: 4x higher<br>65-74 years: 5x higher<br>75-84 years: 8x higher<br>85+ years: 13 x higher | 18 Aug 2020 | COVID-19 Hospitalization and Death by Age [24] | COVID-NET [23] | USA, 13 States |
| Rate ratio for fatality by age | 0-4 years: 9x lower<br>5-17 years: 16x lower<br>18-29: comparison<br>30-39 years: 4x higher<br>40-49 years: 10x higher<br>50-64 years: 30x higher<br>65-74 years: 90x higher<br>75-84 years: 220x higher<br>85+ years: 630x higher | 18 Aug 2020 | COVID-19 Hospitalization and Death by Age [24] | National Center for Health Statistics [25] | USA, 50 States |
| Rate ratio for confirmed case by race / ethnicity | American Indian or Alaska Native, 1.8x;<br>Asian, 0.6x; Black, 1.4x; Latino, 1.7x | 30 Nov 2020 | COVID-19 Hospitalization and Death by Race/Ethnicity [26] |  | USA, 50 States |
| Rate ratio for hospitalization by race / ethnicity | American Indian or Alaska Native, 4.0x;<br>Asian, 1.2x; Black, 3.7x; Latino, 4.1x | 30 Nov 2020 | COVID-19 Hospitalization and Death by Race/Ethnicity [26] | COVID-Net [23] | USA, 13 States |
| Rate ratio for fatality by race / ethnicity | American Indian or Alaska Native, 2.6x;<br>Asian, 1.1x; Black, 2.8x; Latino, 2.8x | 30 Nov 2020 | COVID-19 Hospitalization and Death by Race/Ethnicity [26] | NCHS provisional death counts [27] | USA, 50 States |
| **Disease burden** |  |  |  |  |  |
| Share of ambulatory visits for influenza like illness | 0.7 -3.3% | 2021, week 4 | CovidView [28] | Influenza-like-illness Network [29] | USA |
| Share of emergency department visits for coronavirus-like-illness | 5.1% | 2021, week 4 | CovidView [30] | National Syndromic Surveillance Program (NSSP) [31] | USA, 47 States |
| SARS-CoV-2 laboratory-test-associated hospital admissions | 7.6/100,000 persons | 2021, week 4 | CovidView [32] | COVID-NET [32] | USA, 13 regions |
| Percentage of deaths attributed to pneumonia, influenza, or COVID-19 (PIC) | 28.4% | 2021, week 4 | CovidView [33] | National Center for Vital Statistics [34] | USA, 50 States |
| SARS-CoV-2 antibody seroprevalence | 1.4 – 24% | Dec 2020 | Nationwide Commercial Laboratory | CDC | USA, 49 States |
|  |  |  | Seroprevalence Survey [35] |  |  |
| Cumulative attack rate | 25,412 / 100,000 persons | 19 Jan 2021 | Estimated Disease Burden of COVID-19 [36] | CDC | USA, 50 States |

CDC reports authoritative estimates of most aforementioned infection transmission parameters with the exception of setting-specific secondary infection risks. The majority of CDC transmission data estimates have relied on epidemiological studies conducted outside the U.S.

With regards to disease severity, CDC estimates the asymptomatic fraction as 40% but with a wide range (10-70%) based on summary estimates from meta-analytic reviews as well as individual studies. [1] CDC estimated case fatality and case hospitalization ratios in May 2020 but later removed these parameters. [37] In September 2020, CDC adapted infection fatality ratios from an analysis of European data. [1]

CDC estimates risk ratios for hospitalization and death for age groups and race/ethnicity based on data from the COVID-NET active surveillance program. [38] No estimates of relative risks for infection, hospitalization or death exist for occupations or particular community activities or settings.

Since April 2020, CDC has reported several COVID-19 disease incidence measures on a weekly basis, including emergency department visits for coronavirus-like illness (CLI), SARS-CoV-2 associated hospital admissions, and deaths from pneumonia, influenza, and COVID-19 [39]. CDC additionally reports some of these incidence measures at the level of states or multi-state regions. CLI emergency department visits represent those for fever plus cough and/or shortness of breath or difficulty breathing and come from National Syndromic Surveillance Program (NSSP) that collects and integrates data on ED visit symptoms from a large share of the countries emergency department in 47 states. CDC’s COVID-NET program conducts active surveillance for laboratory-confirmed SARS-CoV-2 hospitalizations in 13 participating sub-state regions publishing age-stratified incidence rates weekly. CDC reports the percentage of deaths attributed to pneumonia or influenza as well as weekly counts of COVID-19 attributed deaths for each state parsed by age, gender, and race / ethnicity. Total excess deaths by cause of death is also available for each state. [40]

In July 2020, CDC reported seroprevalence estimates from 10 sites based on COVID-19 antibodies detected in serum samples collected by commercial laboratories for reasons other than COVID-19. [41] After Aug 2020, CDC expanded seroprevalence reporting to all U.S. states and has been reporting estimates every two weeks. In December 2020, CDC provided the first estimates of cumulative prevalence of infection and symptomatic infection at a national level. [42]

CDC affiliated investigators published 133 epidemiologic studies through Oct 30, 2020. (Supplemental materials Table S3) Of these 133 studies, 99 were descriptive and 34 analytic. Table 3 tabulates study characteristics for both groups of studies.

**Table 3.**
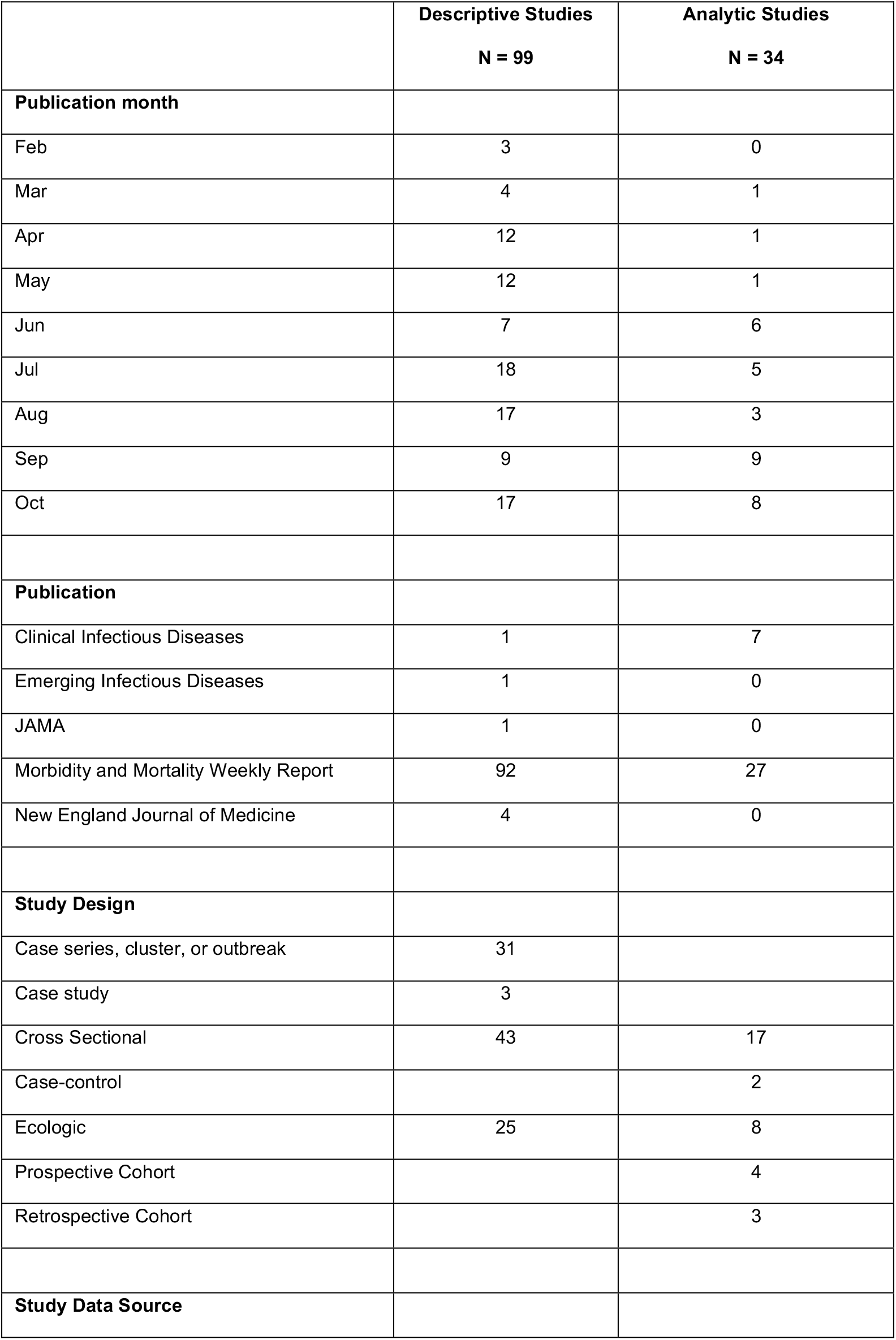
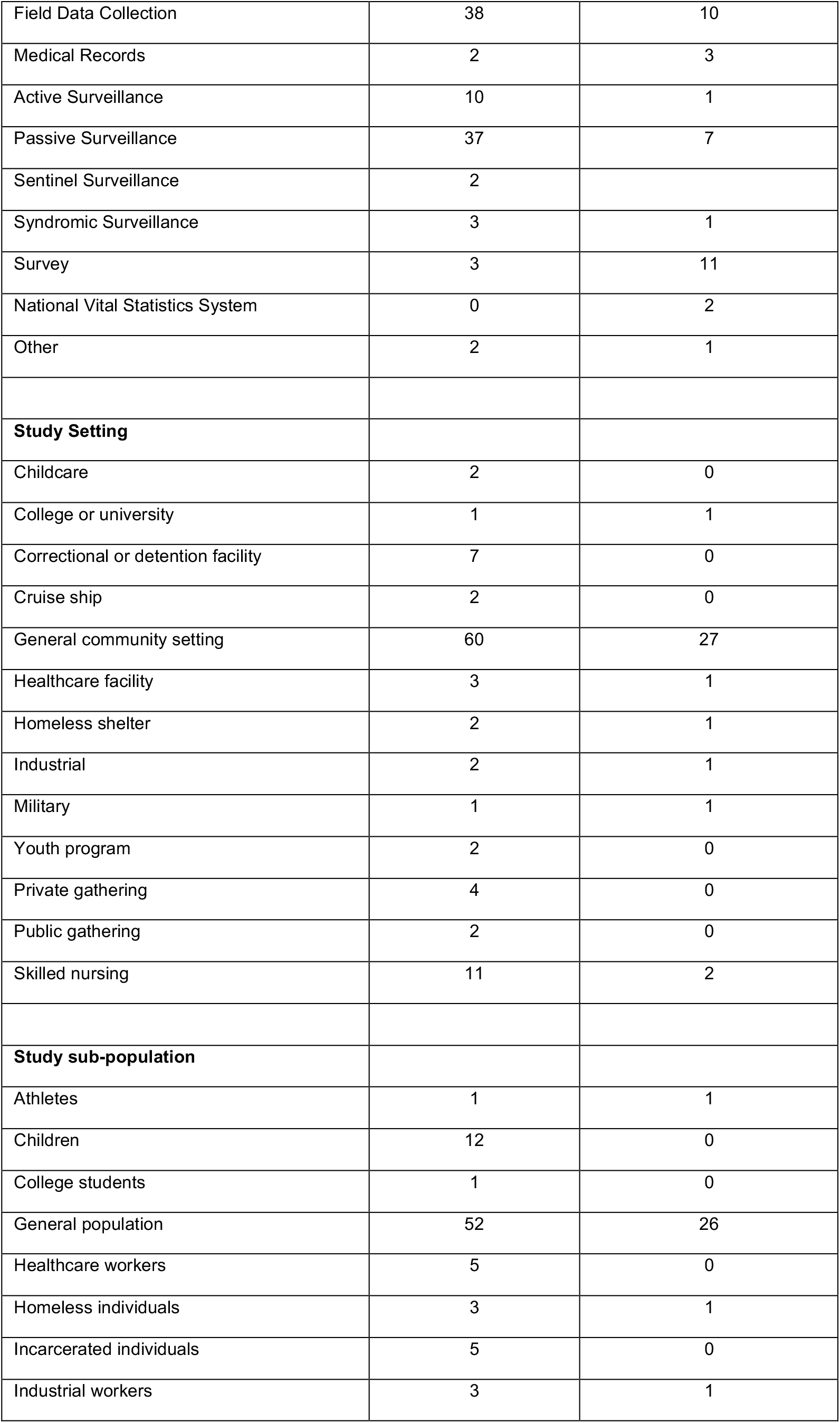
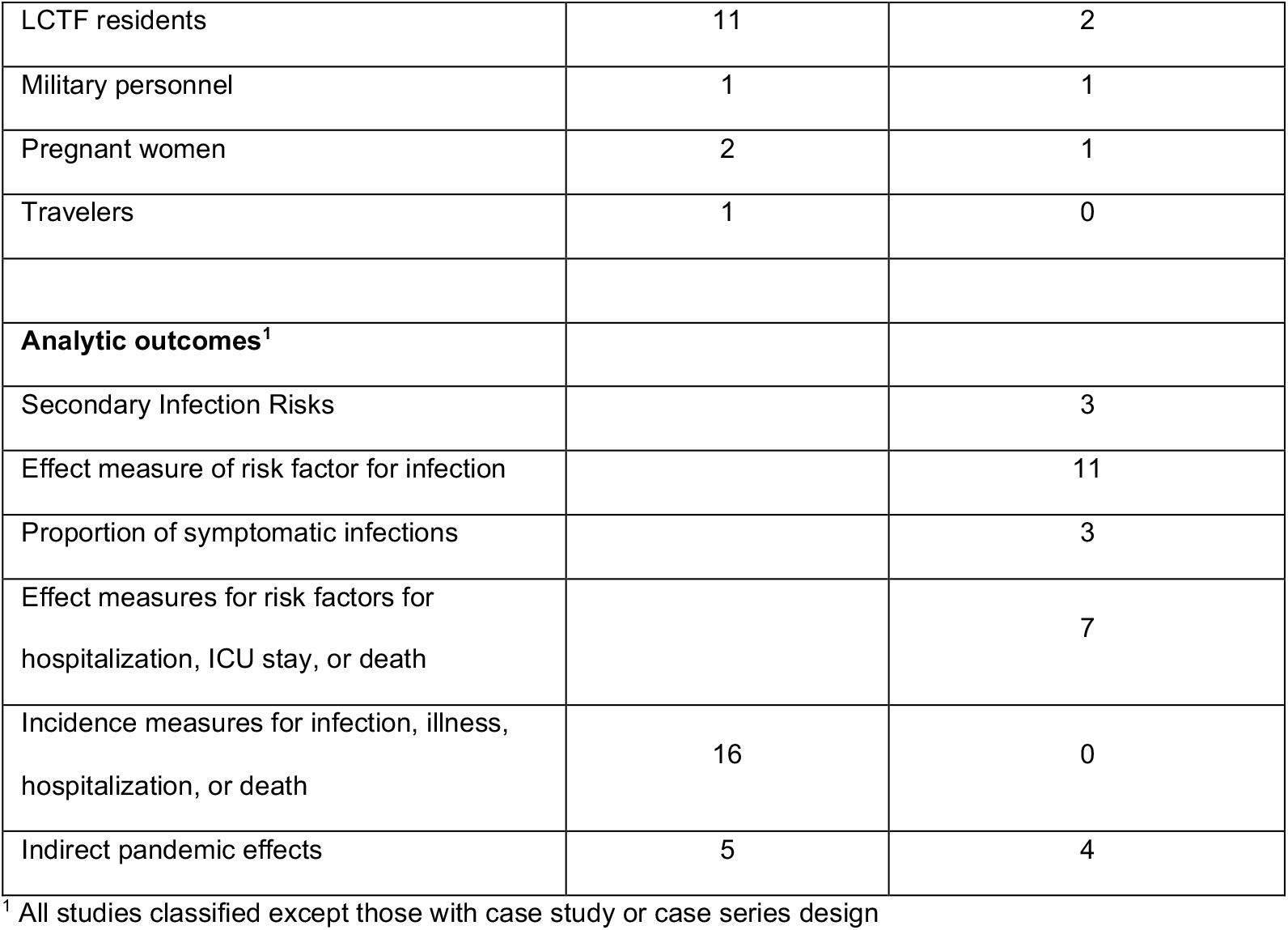
Published epidemiologic studies on COVID-19 transmission, infection severity, and disease burden authored by CDC affiliated investigators

|  | **Descriptive Studies**<br>**N = 99** | **Analytic Studies**<br>**N = 34** |
| --- | --- | --- |
| **Publication month** |  |  |
| Feb | 3 | 0 |
| Mar | 4 | 1 |
| Apr | 12 | 1 |
| May | 12 | 1 |
| Jun | 7 | 6 |
| Jul | 18 | 5 |
| Aug | 17 | 3 |
| Sep | 9 | 9 |
| Oct | 17 | 8 |
| **Publication** |  |  |
| Clinical Infectious Diseases | 1 | 7 |
| Emerging Infectious Diseases | 1 | 0 |
| JAMA | 1 | 0 |
| Morbidity and Mortality Weekly Report | 92 | 27 |
| New England Journal of Medicine | 4 | 0 |
| **Study Design** |  |  |
| Case series, cluster, or outbreak | 31 |  |
| Case study | 3 |  |
| Cross Sectional | 43 | 17 |
| Case-control |  | 2 |
| Ecologic | 25 | 8 |
| Prospective Cohort |  | 4 |
| Retrospective Cohort |  | 3 |
| **Study Data Source** |  |  |
| Field Data Collection | 38 | 10 |
| Medical Records | 2 | 3 |
| Active Surveillance | 10 | 1 |
| Passive Surveillance | 37 | 7 |
| Sentinel Surveillance | 2 |  |
| Syndromic Surveillance | 3 | 1 |
| Survey | 3 | 11 |
| National Vital Statistics System | 0 | 2 |
| Other | 2 | 1 |
| **Study Setting** |  |  |
| Childcare | 2 | 0 |
| College or university | 1 | 1 |
| Correctional or detention facility | 7 | 0 |
| Cruise ship | 2 | 0 |
| General community setting | 60 | 27 |
| Healthcare facility | 3 | 1 |
| Homeless shelter | 2 | 1 |
| Industrial | 2 | 1 |
| Military | 1 | 1 |
| Youth program | 2 | 0 |
| Private gathering | 4 | 0 |
| Public gathering | 2 | 0 |
| Skilled nursing | 11 | 2 |
| **Study sub-population** |  |  |
| Athletes | 1 | 1 |
| Children | 12 | 0 |
| College students | 1 | 0 |
| General population | 52 | 26 |
| Healthcare workers | 5 | 0 |
| Homeless individuals | 3 | 1 |
| Incarcerated individuals | 5 | 0 |
| Industrial workers | 3 | 1 |
| LCTF residents | 11 | 2 |
| Military personnel | 1 | 1 |
| Pregnant women | 2 | 1 |
| Travelers | 1 | 0 |
| **Analytic outcomes<sup>1</sup>** |  |  |
| Secondary Infection Risks |  | 3 |
| Effect measure of risk factor for infection |  | 11 |
| Proportion of symptomatic infections |  | 3 |
| Effect measures for risk factors for hospitalization, ICU stay, or death |  | 7 |
| Incidence measures for infection, illness, hospitalization, or death | 16 | 0 |
| Indirect pandemic effects | 5 | 4 |

<sup>1</sup> All studies classified except those with case study or case series design

Of the 99 descriptive studies we identified, thirty-one were case series, 43 were cross-sectional, and 25 were ecologic. Most descriptive studies summarized laboratory confirmed cases of SARS-CoV-2 infection from a particular setting, disaggregating counts by clinical outcomes, by patient demographic or medical attributes, by exposure type, or by temporal patterns. Descriptive studies reported data from a wide range of populations and community settings.

Some descriptive studies postulated mechanisms of transmission or prevention; however, they did not estimate quantitatively infection transmission or disease severity parameters. Sixteen of the descriptive studies reported on the incidence or prevalence of laboratory confirmed infection, clinical illness, hospitalization or death in either the general population or a defined subpopulation (e.g., skilled nursing facility residents, healthcare workers, incarcerated or homeless individuals) for a defined time period. [43, 44, 45, 46, 47, 41, 48, 49, 50, 51, 52, 53, 54, 55, 56, 57]. One study provided a point estimate of excess deaths associated with COVID-19 using national vital statistics data. [57]

We classified thirty-four studies as analytic. Most (31/34) were published after June 2020 and in the publication MMWR (27/34). The remainder were published in other peer-reviewed journals. Most of the analytic studies had general population subjects (27/34), had cross-sectional (17/34) or ecologic (8/34) designs, and utilized field (10/34) or survey data (11/24).

Most analytic studies estimated at least one parameter for transmission, infection severity, or disease burden. Of the analytic studies estimating transmission parameters, three estimated secondary infection risks (SIR) [58, 59, 60] One of these was a follow up study of the first 12 confirmed cases in the US [58] Two others estimated the SIR for household contacts [59,60]. Our searches found no studies estimating the SIR for workplace, school or community settings or for particular occupations or industries.

Eleven analytic studies provided estimates of an effect measure for risk factors for laboratory-confirmed infections. [59,61, 62, 63, 64, 65, 66, 67, 68, 69, 70] Five of the eleven were conducted using a sample of the general population. These examined the frequency or particular demographics, [61] neighborhood deprivation/social vulnerability levels, [63,68] relationship to a confirmed case, [59] and recent social activities, including restaurant dining. [66] Other investigations examined various factors related to confirmed infection in sub-populations, including face coverings and distancing among occupants of a military ship, [62] ethnic composition of employees of industrial facilities, [64] shelter residence status among people experiencing homelessness, [69] screening strategies and staffing levels in skilled nursing facilities. [65,67] Eight of the eleven studies employed cross-sectional or ecologic designs. No studies examined the independent effects of etiologic variables in multi-variate analysis.

Seven studies examined risk factors for severe disease outcomes, including hospitalization, ICU stay and death. Most of these used general population subjects. [52, 63,71, 72, 73, 74, 75] These studies consistently found older age to be a strong predictor of need for ICU stays, mechanical ventilation and of death. Male gender, end-stage renal disease, coronary artery disease, and neurologic disorders, immunocompromising conditions, pregnancy, African American identity were also correlated with severe disease. [71,72,73,52,75] One study estimated the odds of prolonged symptoms in relation to disease severity. [74]

No CDC-affiliated studies estimated infection-hospitalization or infection-fatality ratios. Three studies assessed symptom prevalence or the fraction of symptomatic people among individuals with positive tests. One study, conducted in a military setting, found that 18.5% (44/238) of those with positive tests were asymptomatic. [62] Two community studies found that nearly all household contacts of an index case had symptoms or developed them shortly after testing positive. [76,77]

Nine analytic studies reported on indirect health impacts of COVID-19, including impacts on emergency departments visits, trends in childhood vaccination, telehealth visits, workplace absenteeism, mental health symptoms, and characteristics associated with delay or avoidance of medical care. [78, 79, 80, 81, 82, 83, 84, 85, 86] One found declines in emergency care visits for heart attacks, strokes, and diabetes; [81] one found an increased prevalence of mental health symptoms; [85] one found increased absenteeism in non-healthcare essential workers; [82] and two documented an initial decline followed by a rebound in childhood vaccination rates. [78,83]

## DISCUSSION

Twelve months after the identification of the SARS-COV-2 virus, epidemiologic understanding of infectious transmission, disease severity, and disease burden has evolved and advanced. However, despite a substantial number of original epidemiological investigations, our review finds data gaps which may limit efficient interventions to prevent COVID-19 transmission.

We restricted this scoping study to epidemiological studies authored or co-authored by CDC investigators. Thus, our review may have also overlooked data found in pre-prints and other unpublished analyses. However, estimating transmission, infection severity and disease burden parameters generally require detailed individual case records or contact tracing data for which access is restricted to government agencies.

### Infectious Transmission

CDC’s current estimates of SARS-CoV-2 transmission parameters, including the incubation period, serial interval, and timing and duration of infectiousness, come from studies conducted outside the U.S. yet are consistent across settings and time-periods and thus considered reliable. On the other hand, CDC estimates of the clinical fraction have a wide range, reflecting significant heterogeneity among study methods, settings and populations. [87,88, 89, 90] Large or multi-regional case-ascertained studies or examination of contact tracing follow up data conducted in the US could provide more precise and age-specific estimates of this parameter as well as of the impact of asymptomatic spread in different settings.

Context-specific estimates of the SIR, including for schools, workplaces, and other community settings, remain unavailable. These parameters require large or setting-specific transmission studies. Available estimates of the SIR for household transmission also vary widely with few based on US settings. No identified transmission studies have quantitatively examined the efficacy of modifying exposure factors within households or workplaces.

Current guidance for COVID-19 community mitigations relies primarily on indirect evidence. For example, pre-symptomatic transmission, proximity, and an inability to mask are the facts used to justify restrictions on indoor dining in restaurants; [91] Such restrictions may well be protective; however, rigorous studies quantifying transmission risks for restaurant settings have not been undertaken.

Studies comparing estimates of the SIR across community settings and activities could identify high risk activities and prioritize risk reduction interventions. Large coordinated case-ascertained prospective studies in particular could be a valuable tool to determine risks from specific activities in household, community, business, transportation, and educational settings and assess the risk reduction associated with operating rules for interrupting transmission. Such studies could inform the relative population-level impacts of social distancing, indoor versus outdoor settings, infectivity of high-touch surfaces, and the value of indoor ventilation. The state-to-state variation in policy response also provides natural experiments to evaluate the relative effectiveness of mandatory and voluntary community mitigations measures.

While several case series and outbreak investigations have highlighted potential hazards in “essential” and frontline work settings such as food production, no identified analytic studies quantified determinants of risks of transmission in workplace settings. Case control studies examining comparative risks across industries and occupations could contribute to understanding how occupational exposures contribute, both directly and through household exposures, to COVID-19 racial disparities.

Studies authored by CDC investigators during the 2009 influenza H1N1 pandemic illustrate the value of timely, robust transmission epidemiology. During 2009, CDC investigators conducted a number of influential transmission studies within the first nine months of the onset of the H1N1 epidemic. One, conducted on the basis of state-submitted case reports in the first months of the epidemic, found that odds ratios for H1N1 infection among household contacts were 3.5 times (1.6 – 8.0) higher for 0-4 year old and 2 times (1.1 - 3.6) higher for 5-18 years old than for 19-50 year old individuals. [92] Another CDC study combined data from 7 household transmission studies early in the 2009 pandemic to quantify the timing of secondary infections within households, demonstrating that about 5% happen >3 days after onset of symptoms. [93] A meta-analysis of 47 studies on transmission rates in households, schools, workplaces, and social events, including fifteen in settings within the U.S., identified multiple population determinants of secondary infection risks, including higher risks for students relative to teachers and for boarding versus day schools. [94]

### Disease Severity

CDC’s estimates of the infection-fatality ratios (IFR) based on studies in six European countries may not be generalizable to the US. County to country differences in the prevalence of vulnerability factors, medical care, and ascertainment of infection affect this parameter. Community cohort studies could provide assessment of multiple infection severity parameters.

While the CDC has established older age as a strong risk factor for severe COVID-19 disease and characterized the relative risk associated with individual medical co-morbidities, [95] further inquiry could examine how the effects of age and co-morbidities vary for individuals with different levels of functional status and in different residential settings (e.g., skilled nursing, assisted living or independent community living).

### Burden of infection and disease

To monitor disease burden, the CDC has routinely reported several measures of infection and disease incidence at the national and state level; however, these measures have limited geographical resolution. Each state has utilized its own distinct sets of state and regional indicators, [96] along with state-specific benchmarks, to guide control activities.

For calibrating mitigations, most states have relied heavily on laboratory-test-based “case” counts, which underestimate infection rates and vary depending on testing access and demand. [97, 6] For such reasons, the US National Pandemic Strategy called for transitioning from counting individual confirmed cases to monitor epidemic trends to monitoring illness rates (i.e., hospitalization admissions and syndromic surveillance) which are generally less subject to bias. [3] In fact, during the H1N1 pandemic, CDC discontinued state reporting of individual lab-confirmed cases two months after the initiation of the epidemic and initiated state reporting of total numbers of weekly H1N1 hospitalizations and deaths. [5] The CDC used these numbers along other surveillance data to provide monthly estimates of symptomatic illness, hospitalizations, and deaths. [98] During H1N1, CDC also used syndromic surveillance data to model infection dynamics at sub-state regional level, illustrating important regional and temporal patterns of transmission relevant for control and planning decisions. [5]

A robust, national approach to monitoring COVID-19 in every region using multiple measures of disease incidence should be feasible. In July 2020, the US Department of Health and Human Services required all hospitals to report COVID-19 associated admissions to the Federal government as part of a new national reporting standard; this data should allow publishing real time estimates of new hospital admissions by age at a regional level. Coronavirus-like-illness emergency department visits from the NSSP could also be a nationally consistent regional measure. Consistent application of surveillance measures across U.S. regions could lead to better informed and targeted mitigations and their evaluation.

Disaggregating measures of infection and disease burden by community setting (e.g. residential setting, workplaces, neighborhoods) could further aide in targeting public health responses and focusing resources for testing, contact tracing and outbreak investigations. Since May, CMS has required reporting of confirmed cases and deaths from regulated skilled nursing facilities; however, there is still little systematic data on the burden of infection, morbidity, and mortality in other community settings. Through the National Notifiable Disease Surveillance System (NNDSS), CDC has accumulated over 20 million case reports. These reports should contain exposure information on residence, occupation, travel. Analytic studies could combine exposure prevalence among confirmed cases along with population exposure prevalence to estimate attributable risks for each exposure.

The US currently lacks regional estimates of the cumulative infection prevalence. Regional differences in prevalence could explain part of the variation in the pace of infectious spread. Methods to assess exposure using antibodies have evolved and improved over the epidemic time course. [99,100] Applying serial seroprevalence studies both to estimate cumulative incidence and infection fatality rates should be an important subject of investigation.

### Conclusions

Twelve months after the identification of the SARS-CoV-2 virus, the understanding of COVID-19 transmission, infection severity and disease burden has advanced, yet estimates of several essential epidemiological parameters remain absent or uncertain. Missing are comparative measures of transmission risk and disease burden for community exposure settings, including work in “essential occupations.” Estimates for infection fatality and infection hospitalization ratios representative of US settings do not exist. Indicators of disease burden, though available, have insufficient resolution to inform targeted policy and programmatic responses.

These epidemiological data gaps may be limiting the most efficient and equitable response to the COVID-19 epidemic and underscore the importance of standardizing data collection priorities and protocols early during a rapidly emerging infectious disease epidemic. CDC scientists and staff have authored a large number of studies on COVID-19; however, the majority are descriptive and provide low-quality evidence for policy and management decisions. The content of their investigations raises questions about whether and how an explicit national research agenda guided CDC epidemiological endeavors.

CDC scientists have the access to data, the expertise, and the resources to provide the data necessary for an optimal epidemic response. Moving forward, the CDC should now plan for how it might develop and implement a timely, strategic, and prioritized national epidemiological data collection and research agenda for the next emerging infectious disease epidemic.

## Data Availability

Data used for this study are included in supplemental materials and are additionaly publicly available

