## Supplemental Tables for "The Missing Science: Epidemiological data gaps for COVID-19 policy in the United States"

### Table S1. Infection transmissibility and severity parameters for Influenza H1N1 (2009) within the Pandemic Severity Assessment Framework

| Parameter | Range of anticipated values | Value for Influenza H1N1 | Severity score (1 = Least severe to 7 = most severe) |
|---|---|---|---|
| **Transmissibility** | | | |
| Symptomatic attack rate, community, % | <10 % to >25% | 20% | 3 |
| Symptomatic attack rate, school or university, % | <20% to >36% | NA | NA |
| Symptomatic attack rate, workplace, % | <10 % to >25% | NA | NA |
| Household secondary attack rate, symptomatic, % | < 5% to > 21% | 13% | 3 |
| R0: reproductive number | <1.1 to >1.8 | 1.4 | 3 |
| Peak % outpatient illness visits | 1-3 % to >13% | 7% | 3 |
| **Clinical severity** | | | |
| Case-fatality ratio (Infection fatality ratio) | < 0.02 to > 1% | 0.02% | 2 |
| Case-hospitalization ratio | < 0.5 to > 7% | 0.05% | 2 |
| Ratio, deaths: hospitalizations | <3 to > 18% | 4.7% | 2 |

### Table S2. Accessed Centers for Disease Control Webpages

| Webpage | Version Date | Current Website URL |
|---|---|---|
| COVID-19 Pandemic Planning Scenarios | 10 September 2020 | https://www.cdc.gov/coronavirus/2019-ncov/hcp/planning-scenarios.html |
| COVID-19 Hospitalization and Death by Age | 18 Aug 2020 | https://www.cdc.gov/coronavirus/2019-ncov/covid-data/investigations-discovery/hospitalization-death-by-age.html |
| COVID-19 Hospitalization and Death by Race/Ethnicity | 30 Nov 2020 | https://www.cdc.gov/coronavirus/2019-ncov/covid-data/investigations-discovery/hospitalization-death-by-race-ethnicity.html |
| COVIDView: A weekly surveillance summary of COVID-19 activity in the U.S. | Updated Weekly | https://www.cdc.gov/coronavirus/2019-ncov/covid-data/covidview/index.html |
| Estimated Disease Burden of COVID-19 | 3 Dec 2021 | https://www.cdc.gov/coronavirus/2019-ncov/cases-updates/burden.html |
| Commercial Laboratory Seroprevalence Survey Data | 21 July 2020 | https://covid.cdc.gov/covid-data-tracker/#serology-surveillance |
| Nationwide Commercial Laboratory Seroprevalence Survey | Updated bi-monthly | https://covid.cdc.gov/covid-data-tracker/#national-lab |
| Coronavirus Disease 2019 (COVID-19)-Associated Hospitalization Surveillance Network (COVID-NET) | Updated Weekly | https://www.cdc.gov/coronavirus/2019-ncov/covid-data/covid-net/purpose-methods.html |
| United States COVID-19 Cases and Deaths by State | Updated Daily | https://covid.cdc.gov/covid-data-tracker/#cases_casesper100klast7days |
| Evidence used to update the list of underlying medical conditions that increase a person's risk of severe illness from COVID-19 | 2 Nov 2020 | https://www.cdc.gov/coronavirus/2019-ncov/need-extra-precautions/evidence-table.html |

Table S3. List of reviewed CDC-affiliated studies.

| Month | First author | Title | Publication | DOI | Category | Study category | Data source | Description of study outcomes | Setting | Sub-population | Secondary Infection Risk | Effect measures of any risk factor for infection | Effect measures for any risk factors for hospitalization, ICU stay, or death | Symptom prevalence or symptomatic infection fraction | Infection hospitalization or fatality ratios | Measure of incidence or prevalence of infection, clinical illness, hospitalization, or death | Any epidemiologic measure of an indirect pandemic effect |
|---|---|---|---|---|---|---|---|---|---|---|---|---|---|---|---|---|---|
| February | Patel | Initial Public Health Response and Interim Clinical Guidance for the 2019 Novel Coronavirus Outbreak — United States, December 31, 2019–February 4, 2020 | MMWR | http://dx.doi.org/10.15585/mmwr.mm6905e1external icon | Descriptive | Case series or cluster | Passive surveillance program | Frequency of exposure characteristics of initial 11 U.S. 2019 novel coronavirus cases | General community | General population | | | | | | | |
| February | Bajema | Persons Evaluated for 2019 Novel Coronavirus — United States, January 2020 | MMWR | http://dx.doi.org/10.15585/mmwr.mm6906e1 | Descriptive | Case series or cluster | Passive surveillance program | Frequency of clinical characteristics and epidemiologic risk factors persons tested for 2019 novel coronavirus | General community | General population | | | | | | | |
| February | Jernigan | Update: Public Health Response to the Coronavirus Disease 2019 Outbreak — United States, February 24, 2020 | MMWR | http://dx.doi.org/10.15585/mmwr.mm6908e1 | Descriptive | Case series or cluster | Passive surveillance program | Frequency of exposure characteristics of initial 14 U.S. 2019 novel coronavirus cases diagnosed withing the U.S. | General community | General population | | | | | | | |
| March | Burke | Active Monitoring of Persons Exposed to Patients with Confirmed COVID-19 — United States, January–February 2020 | MMWR | http://dx.doi.org/10.15585/mmwr.mm6909e1 | Analytic | Prospective cohort | Field data collection | Secondary infection risks for household and non-household contacts | General community | General population | X | | | | | | |
| March | McMichael | COVID-19 in a Long-Term Care Facility — King County, Washington, February 27–March 9, 2020 | MMWR | http://dx.doi.org/10.15585/mmwr.mm6912e1external icon | Descriptive | Case series or cluster | Field data collection | Frequency of characteristics of patients with COVID-19 epidemiologically linked to skilled nursing facility outbreak | Skilled nursing facilities | LCTF residents | | | | | | | |
| March | ovid-19 Respons | Severe Outcomes Among Patients with Coronavirus Disease 2019 (COVID-19) — United States, February 12–March 16, 2020 | MMWR | http://dx.doi.org/10.15585/mmwr.mm6912e2external icon | Descriptive | Cross-sectional | Passive surveillance program | Frequency of clinical outcomes among initial 4226 confirmed U.S. novel coronavirus cases | General community | General population | | | | | | | |
| March | Moriarty | Public Health Responses to COVID-19 Outbreaks on Cruise Ships — Worldwide, February–March 2020 | MMWR | http://dx.doi.org/10.15585/mmwr.mm6912e3 | Descriptive | Ecologic | Field data collection | Frequency of demographic characteristics of passengers and crew members on board two cruise ships with COVID-19 outbreaks | Cruise ship | Travelers | | | | | | | |
| March | McMichael | Epidemiology of Covid-19 in a Long-Term Care Facility in King County, Washington | NEJM | https://www.nejm.org/doi/full/10.1056/NEJMoa2005412 | Descriptive | Case series or cluster | Field data collection | Frequency of demographic and Clinical Characteristics of Persons with Confirmed Covid-19 Linked to skilled nursing facility exposure | Skilled nursing facilities | Elderly | | | | | | | |
| April | ovid-19 Respons | Preliminary Estimates of the Prevalence of Selected Underlying Health Conditions Among Patients with Coronavirus Disease 2019 — United States, February 12–March 28, 2020 | MMWR | http://dx.doi.org/10.15585/mmwr.mm6913e2external icon | Analytic | Cross-sectional | Passive surveillance program | Frequency of clinical outcomes and hospitalization status among COVID-19 patients by demographic and clinical characteristics | General community | General population | | | X | | | | |
| April | Kimball | Asymptomatic and Presymptomatic SARS-CoV-2 Infections in Residents of a Long-Term Care Skilled Nursing Facility — King County, Washington, March 2020 | MMWR | http://dx.doi.org/10.15585/mmwr.mm6913e1 | Descriptive | Case series or cluster | Field data collection | Frequency of demographic characteristics and reported symptoms among skilled nursing facility residents by SARS-Cov-test status | Skilled nursing facilities | LCTF residents | | | | | | | |
| April | Roxby | Detection of SARS-CoV-2 Among Residents and Staff Members of an Independent and Assisted Living Community for Older Adults — Seattle, Washington, 2020 | MMWR | http://dx.doi.org/10.15585/mmwr.mm6914e2external icon | Descriptive | Case series or cluster | Field data collection | Frequency of characteristics of skilled nursing facility residents and staff members with positive SARS-CoV-2 test results | Skilled nursing facilities | LCTF residents | | | | | | | |
| April | Zwald | Rapid Sentinel Surveillance for COVID-19 — Santa Clara County, California, March 2020 | MMWR | http://dx.doi.org/10.15585/mmwr.mm6914e3external icon | Descriptive | Cross-sectional | Sentinel surveillance program | Number of positive test results for SARS-Cov-2 among respiratory specimins submitted for influenza | General community | General population | | | | | | | |
| April | ovid-19 Respons | Coronavirus Disease 2019 in Children — United States, February 12–April 2, 2020 | MMWR | http://dx.doi.org/10.15585/mmwr.mm6914e4 | Descriptive | Cross-sectional | Passive surveillance program | Frequency of signs and symptoms among pediatric and adult patients with laboratory-confirmed COVID-19 | General community | Children | | | | | | | |
| April | ovid-19 Respons | Geographic Differences in COVID-19 Cases, Deaths, and Incidence — United States, February 12–April 7, 2020 | MMWR | http://dx.doi.org/10.15585/mmwr.mm6915e4 | Descriptive | Ecologic | Passive surveillance program | Cumulative number of reported COVID-19 cases, by jurisdiction | General community | General population | | | | | | | |

Table S3. List of reviewed CDC-affiliated studies.

| | | | | | | | | | | | | | |
|---|---|---|---|---|---|---|---|---|---|---|---|---|---|
| April | Ghinai | Community Transmission of SARS-CoV-2 at Two Family Gatherings — Chicago, Illinois, February–March 2020 | MMWR | http://dx.doi.org/10.15585/mmwr.mm6915e1 | Descriptive | Case series or cluster | Field data collection | Incubation periods | Private social gathering | General population | | | |
| April | Heinzerling | Transmission of COVID-19 to Health Care Personnel During Exposures to a Hospitalized Patient — Solano County, California, February 2020 | MMWR | http://dx.doi.org/10.15585/mmwr.mm6915e5external icon | Descriptive | Case series or cluster | Field data collection | Frequency of demographic and exposure risk characteristics health care workers exposed to an index case | Healthcare facility | Healthcare workers | | | |
| April | ›vid-19 Respons | Characteristics of Health Care Personnel with COVID-19 — United States, February 12–April 9, 2020 | MMWR | http://dx.doi.org/10.15585/mmwr.mm6915e6 | Descriptive | Cross-sectional | Passive surveillance program | Frequency of demographic characteristics, exposures, symptoms, and underlying health conditions among health care personnel with COVID-19 | General community | Healthcare workers | | | |
| April | Lasry | Timing of Community Mitigation and Changes in Reported COVID-19 and Community Mobility — Four U.S. Metropolitan Areas, February 26–April 1, 2020 | MMWR | http://dx.doi.org/10.15585/mmwr.mm6915e2 | Descriptive | Ecologic | Passive surveillance program | TIming of community mitigation measures relative to COVID-19 confirmed case counts | General community | General population | | | |
| April | Garg | Hospitalization Rates and Characteristics of Patients Hospitalized with Laboratory-Confirmed Coronavirus Disease 2019 — COVID-NET, 14 States, March 1–30, 2020 | MMWR | http://dx.doi.org/10.15585/mmwr.mm6915e3external icon | Descriptive | Cross-sectional | Active surveillance program | Laboratory-confirmed coronavirus disease 2019 (COVID-19)–associated hospitalization incidence rates by age group and surveillance site | General community | General population | | × | |
| April | Chang | Cleaning and Disinfectant Chemical Exposures and Temporal Associations with COVID-19 — National Poison Data System, United States, January 1, 2020–March 31, 2020 | MMWR | http://dx.doi.org/10.15585/mmwr.mm6916e1external icon | Descriptive | Cross-sectional | Passive surveillance program | Trends in poison control reports for exposures to cleaners and disinfectants | General community | General population | | | |
| April | Arons | Presymptomatic SARS-CoV-2 Infections and Transmission in a Skilled Nursing Facility | NEJM | https://www.nejm.org/doi/full/10.1056/NEJMoa2008457 | Descriptive | Case series or cluster | Field data collection | Frequency of demographic Characteristics and reported Symptoms among skilled nursing facility residents by SARS-Cov- test status | Skilled nursing facilities | LCTF residents | | | |
| May | Tobolowsky | COVID-19 Outbreak Among Three Affiliated Homeless Service Sites — King County, Washington, 2020 | MMWR | http://dx.doi.org/10.15585/mmwr.mm6917e2external icon | Descriptive | Case series or cluster | Field data collection | Prevalence of positive SARS-CoV-2 tests | Homeless shelter | Homeless individuals | | | |
| May | Mosites | Assessment of SARS-CoV-2 Infection Prevalence in Homeless Shelters — Four U.S. Cities, March 27–April 15, 2020 | MMWR | http://dx.doi.org/10.15585/mmwr.mm6917e1 | Descriptive | Ecologic | Field data collection | Prevalence of positive SARS-CoV-2 tests | Homeless shelter | Homeless individuals | | × | |
| May | Gold | Characteristics and Clinical Outcomes of Adult Patients Hospitalized with COVID-19 — Georgia, March 2020 | MMWR | http://dx.doi.org/10.15585/mmwr.mm6918e1 | Descriptive | Cross-sectional | Medical records | Frequency of medical conditions among adults hospitalized with COVID-19 by age group and race/ethnicity | General community | General population | | | |
| May | Rosenberg | New York State Coronavirus 2019 Response Team. COVID-19 Testing, Epidemic Features, Hospital Outcomes, and Household Prevalence, New York State-March 2020 | CID | 10.1093/cid/ciaa549 | Analytic | Cross-sectional | Passive surveillance program | Frequency of demographics, Risk Factors, Comorbidities, Symptoms, and Hospital Outcomes for Persons Diagnosed With Coronavirus Disease 2019; household attack rate | General community | General population | × | | |
| May | Wallace | COVID-19 in Correctional and Detention Facilities — United States, February–April 2020 | MMWR | http://dx.doi.org/10.15585/mmwr.mm6919e1external icon | Descriptive | Cross-sectional | Passive surveillance program | Frequency of COVID-19 infections, hospitalizations, and deaths among correctional and detention facility residents and staff | Correctional or detention facilties | Incarcerated individuals | | | |
| May | Wallace | Public Health Response to COVID-19 Cases in Correctional and Detention Facilities — Louisiana, March–April 2020 | MMWR | http://dx.doi.org/10.15585/mmwr.mm6919e3external icon | Descriptive | Ecologic | Passive surveillance program | Frequency of characteristics of correctional facilities by presence of COVID-19 cases among incarcerated | Correctional or detention facilties | Incarcerated individuals | | | |
| May | Hamner | High SARS-CoV-2 Attack Rate Following Exposure at a Choir Practice — Skagit County, Washington, March 2020 | MMWR | http://dx.doi.org/10.15585/mmwr.mm6919e6external icon | Descriptive | Case series or cluster | Field data collection | Number of event participants with and without COVID-19–compatible symptoms | Private social gathering | General population | | | |
| May | Santoli | Effects of the COVID-19 Pandemic on Routine Pediatric Vaccine Ordering and Administration — United States, 2020 | MMWR | http://dx.doi.org/10.15585/mmwr.mm6919e2 | Descriptive | Ecologic | Other | Weekly trends in vaccine orders | General community | Children | | | × |
| May | ›vid-19 Respons | Preliminary Estimate of Excess Mortality During the COVID-19 Outbreak — New York City, March 11–May 2, 2020 | MMWR | http://dx.doi.org/10.15585/mmwr.mm6919e5external icon | Descriptive | Ecologic | Passive surveillance program | Number of laboratory-confirmed and probable COVID-19–associated deaths and total estimated excess deaths | General community | General population | | × | |
| May | Hamner | High COVID-19 Attack Rate Among Attendees at Events at a Church — Arkansas, March 2020 | MMWR | http://dx.doi.org/10.15585/mmwr.mm6919e6external icon | Descriptive | Case series or cluster | Field data collection | Frequency of demographic characteristics by church attendance and and SARS-CoV-2 testing status; attack rate by demographic characteristic and church activity | Public social gathering | General population | | | |
| May | Bramer | Decline in Child Vaccination Coverage During the COVID-19 Pandemic — Michigan Care Improvement Registry, May 2016–May 2020 | MMWR | http://dx.doi.org/10.15585/mmwr.mm6920e1 | Descriptive | Ecologic | Passive surveillance program | Trends in the proportion of Michigan infants and children with timely vaccination | General community | Children | | | × |

Table S3. List of reviewed CDC-affiliated studies.

| | | | | | | | | | | | | | | | | |
|---|---|---|---|---|---|---|---|---|---|---|---|---|---|---|---|---|
| May | Dora | Universal and Serial Laboratory Testing for SARS-CoV-2 at a Long-Term Care Skilled Nursing Facility for Veterans — Los Angeles, California, 2020 | MMWR | http://dx.doi.org/10.15585/mmwr.mm6921e1external icon. | Descriptive | Case series or cluster | Field data collection | Frequency of characteristics of long-term care skilled nursing facility residents with positive test results for SARS-CoV-2 | Skilled nursing facilities | LCTF residents | | | | | | |
| May | Chu | Investigation and Serologic Follow-Up of Contacts of an Early Confirmed Case-Patient with COVID-19, Washington, USA | EID | https://dx.doi.org/10.3201/eid2608.201423 | Descriptive | Case series or cluster | Field data collection | Prevalence of positive SARS-CoV-2 tests among contacts | General community | General population | | | | | | |
| June | Marcus | COVID-19 Monitoring and Response Among U.S. Air Force Basic Military Trainees — Texas, March–April 2020 | MMWR | http://dx.doi.org/10.15585/mmwr.mm6922e2 | Descriptive | Cross-sectional | Passive surveillance program | Cumulative number of tested trainees with respiratory symptoms and positive test results for SARS-CoV-2 | Military setting | Military personnel | | | | | | |
| June | Jorden | Evidence for Limited Early Spread of COVID-19 Within the United States, January–February 2020 | MMWR | http://dx.doi.org/10.15585/mmwr.mm6922e1 | Descriptive | Cross-sectional | Syndromic surveillance program | Proportion of emergency department (ED) visits for COVID-19–like illness | General community | General population | | | | | × | |
| June | Payne | SARS-CoV-2 Infections and Serologic Responses from a Sample of U.S. Navy Service Members — USS Theodore Roosevelt, April 2020 | MMWR | http://dx.doi.org/10.15585/mmwr.mm6923e4external fraction | Analytic | Cross-sectional | Field data collection | Frequency / odds of SARS-CoV-2 infection by demographic and clinical characteristics and prevention behaviors; clinical fraction | Military setting | Military personnel | × | | × | | | |
| June | Hartnett | Impact of the COVID-19 Pandemic on Emergency Department Visits — United States, January 1, 2019–May 30, 2020 | MMWR | http://dx.doi.org/10.15585/mmwr.mm6923e1 | Analytic | Ecologic | Syndromic surveillance program | Differences in mean weekly numbers of emergency department (ED) visits by diagnostic categories | General community | General population | | | | | | × |
| June | Gharpure | Knowledge and Practices Regarding Safe Household Cleaning and Disinfection for COVID-19 Prevention — United States, May 2020 | MMWR | http://dx.doi.org/10.15585/mmwr.mm6923e2external icon | Descriptive | Cross-sectional | Survey | Prevalence of cleaning and disinfection practices to prevent SARS-CoV-2 infection | General community | General population | | | | | | |
| June | Stokes | Coronavirus Disease 2019 Case Surveillance — United States, January 22–May 30, 2020 | MMWR | http://dx.doi.org/10.15585/mmwr.mm6924e2external icon | Descriptive | Cross-sectional | Passive surveillance program | Cumulative incidence of reported laboratory-confirmed COVID-19 cases by age; frequency of clinical characteristics of confirmed cases by age | General community | General population | | | | | | |
| June | Czeisler | Public Attitudes, Behaviors, and Beliefs Related to COVID-19, Stay-at-Home Orders, Nonessential Business Closures, and Public Health Guidance — United States, New York City, and Los Angeles, May 5–12, 2020 | MMWR | http://dx.doi.org/10.15585/mmwr.mm6924e1 | Analytic | Cross-sectional | Survey | Prevalence of attitudes, behaviors, and beliefs related to COVID-19, stay-at-home orders, nonessential business closures, and public health guidance, by respondent characteristics | General community | General population | | | | | | |
| June | Dawson | Loss of Taste and Smell as Distinguishing Symptoms of COVID-19 | CID | https://doi.org/10.1093/cid/ciaa799 | Analytic | Cross-sectional | Field data collection | Prevalence of self reported symptoms | General community | General population | | | × | | | |
| June | Ellington | Characteristics of Women of Reproductive Age with Laboratory-Confirmed SARS-CoV-2 Infection by Pregnancy Status — United States, January 20–June 7, 2020 | MMWR | http://dx.doi.org/10.15585/mmwr.mm6925a1 | Analytic | Cross-sectional | Passive surveillance program | Frequency of clinical characteristics; relative risk of hospitalizations, intensive care unit (ICU) admissions, receipt of mechanical ventilation, and deaths among women with laboratory-confirmed SARS-CoV-2 infection by pregnancy status, age group, and race/ethnicity; risk ratios for selected factors | General community | Pregnant women | | × | | | | |
| June | Killerby | Characteristics Associated with Hospitalization Among Patients with COVID-19 — Metropolitan Atlanta, Georgia, March – April 2020 | MMWR | http://dx.doi.org/10.15585/mmwr.mm6925e1external icon | Analytic | Cross-sectional | Medical records | Odds ratios for risk factors for COVID-19 hospitalization | General community | General population | | × | | | | |
| June | Lange | Potential Indirect Effects of the COVID-19 Pandemic on Use of Emergency Departments for Acute Life-Threatening Conditions — United States, January–May 2020 | MMWR | http://dx.doi.org/10.15585/mmwr.mm6925e2external icon | Descriptive | Cross-sectional | Syndromic surveillance program | Percent change in the number of emergency department visits for myocardial infarction, stroke, and hyperglycemic crisis | General community | General population | | | | | | × |
| June | Dufort | Multisystem Inflammatory Syndrome in Children in New York State | NEJM | https://www.nejm.org/doi/full/10.1056/NEJMoa2021756 | Descriptive | Cross-sectional | Active surveillance program | Frequency of demographic and clinical characteristics of children meeting surveillance criteria or MIS-C | General community | Children | | | | | | |
| June | Feldstein | Multisystem Inflammatory Syndrome in U.S. Children and Adolescents | NEJM | https://www.nejm.org/doi/full/10.1056/NEJMoa2021680 | Descriptive | Cross-sectional | Sentinel surveillance program | Frequency of demographic and clinical characteristics of children meeting surveillance criteria or MIS-C | General community | Children | | | | | | |
| July | Lewis | COVID-19 Outbreak Among College Students After a Spring Break Trip to Mexico — Austin, Texas, March 26–April 5, 2020 | MMWR | http://dx.doi.org/10.15585/mmwr.mm6926e1external icon | Descriptive | Case series or cluster | Field data collection | Frequency of symptoms among contacts by RT-PCR test status; calculated odds ratios | General community | College students | | | | | | |
| July | Njunga | Serial Laboratory Testing for SARS-CoV-2 Infection Among Incarcerated and Detained Persons in a Correctional and Detention Facility — Louisiana, April–May 2020 | MMWR | http://dx.doi.org/10.15585/mmwr.mm6926e2external icon | Descriptive | Ecologic | Field data collection | Frequency of reported symptoms among incarcerated persons by SARS-CoV-2 test results | Correctional or detention facilities | Incarcerated individuals | | | | | | |

Table S3. List of reviewed CDC-affiliated studies.

| | | | | | | | | | | | | | | | |
|---|---|---|---|---|---|---|---|---|---|---|---|---|---|---|---|
| July | Tenforde | Characteristics of Adult Outpatients and Inpatients with COVID-19 — 11 Academic Medical Centers, United States, March–May 2020 | MMWR | http://dx.doi.org/10.15585/mmwr.mm6926e3external icon | Descriptive | Cross-sectional | Survey | Frequency of demographic, clinical, symptoms, and exposure characteristics of SARS-CoV-2 RT-PCR test positive inidviduals by inpatient status | General community | General population | | | | | |
| July | Marshall | Exposures Before Issuance of Stay-at-Home Orders Among Persons with Laboratory-Confirmed COVID-19 — Colorado, March 2020 | MMWR | http://dx.doi.org/10.15585/mmwr.mm6926e4 | Descriptive | Cross-sectional | Passive surveillance program | Frequency of contact relationships and settings among SARS-CoV-2 RT-PCR test positive inidviduals before stay at home orders | General community | General population | | | | | |
| July | Callaghan | Screening for SARS-CoV-2 Infection Within a Psychiatric Hospital and Considerations for Limiting Transmission Within Residential Psychiatric Facilities — Wyoming, 2020 | MMWR | http://dx.doi.org/10.15585/mmwr.mm6926a4 | Descriptive | Case series or cluster | Field data collection | Frequency of characteristics of patients and health care personnel | Correctional or detention facilties | LCTF residents | | | | | |
| July | Groenewald | Increases in Health-Related Workplace Absenteeism Among Workers in Essential Critical Infrastructure Occupations During the COVID-19 Pandemic — United States, March–April 2020 | MMWR | http://dx.doi.org/10.15585/mmwr.mm6927a1 | Analytic | Cross-sectional | Survey | Prevalence of health-related workplace absenteeism | General community | General population | | | | | × |
| July | Waltenberg | Update: COVID-19 Among Workers in Meat and Poultry Processing Facilities — United States, April–May 2020 | MMWR | http://dx.doi.org/10.15585/mmwr.mm6927e2 | Descriptive | Cross-sectional | Passive surveillance program | Number of laboratory-confirmed COVID-19 cases among workers in meat and poultry facilities | General community | Industrial workers | | | | | |
| July | Sanchez | Initial and Repeated Point Prevalence Surveys to Inform SARS-CoV-2 Infection Prevention in 26 Skilled Nursing Facilities — Detroit, Michigan, March–May 2020 | MMWR | http://dx.doi.org/10.15585/mmwr.mm6927e1external icon | Descriptive | Ecologic | Field data collection | Serial point prevalence of infection | Skilled nursing facilities | LCTF residents | | | | | |
| July | Hsu | Race/Ethnicity, Underlying Medical Conditions, Homelessness, and Hospitalization Status of Adult Patients with COVID-19 at an Urban Safety-Net Medical Center — Boston, Massachusetts, 2020 | MMWR | http://dx.doi.org/10.15585/mmwr.mm6927a3external icon | Descriptive | Cross-sectional | Medical records | Frequency of demographic and clinical characteristics of inpatients with COVID-19 | General community | Homeless individuals | | | | | |
| July | Kim | Risk Factors for Intensive Care Unit Admission and In-hospital Mortality Among Hospitalized Adults Identified through the US Coronavirus Disease 2019 (COVID-19)-Associated Hospitalization Surveillance Network (COVID-NET) | CID | https://doi.org/10.1093/cid/ciaa1012 | Analytic | Retrospective cohort | Active surveillance program | Odds of ICU admission for age and selected medical co-morbidities | General community | General population | × | | | | |
| July | Hendrix | Absence of Apparent Transmission of SARS-CoV-2 from Two Stylists After Exposure at a Hair Salon with a Universal Face Covering Policy — Springfield, Missouri, May 2020 | MMWR | http://dx.doi.org/10.15585/mmwr.mm6928e2 | Descriptive | Case series or cluster | Field data collection | Frequency of symptoms and positive SARS-CoV-2 tests among a convenience sample | General community | General population | | | | | |
| July | Wortham | Characteristics of Persons Who Died with COVID-19 — United States, February 12–May 18, 2020 | MMWR | http://dx.doi.org/10.15585/mmwr.mm6928e1 | Descriptive | Cross-sectional | Passive surveillance program | Frequency of co-morbid conditions among COVID-19 associated deaths by age group | General community | General population | | | | | |
| July | Burke | Symptom Profiles of a Convenience Sample of Patients with COVID-19 — United States, January–April 2020 | MMWR | http://dx.doi.org/10.15585/mmwr.mm6928a2external icon | Descriptive | Cross-sectional | Field data collection | Frequency of reported symptoms among patients with laboratory-confirmed COVID-19, by age and hospitalization status | General community | General population | | | | | |
| July | Bushman | Detection and Genetic Characterization of Community-Based SARS-CoV-2 Infections — New York City, March 2020 | MMWR | http://dx.doi.org/10.15585/mmwr.mm6928a5external icon | Descriptive | Ecologic | Syndromic surveillance program | Trends in the daily percentage of emergency department (ED) visits for influenza-like illness (ILI), number of confirmed COVID-19 cases, and number and percentage of sentinel specimens positive for SARS-CoV-2 | General community | General population | | | | × | |
| July | Fisher | Factors Associated with Cloth Face Covering Use Among Adults During the COVID-19 Pandemic — United States, April and May 2020 | MMWR | http://dx.doi.org/10.15585/mmwr.mm6928e3external icon | Descriptive | Cross-sectional | Survey | Period prelence of cloth face covering use | General community | General population | | | | | |
| July | Havers | Seroprevalence of Antibodies to SARS-CoV-2 in 10 Sites in the United States, March 23-May 12, 2020 | JAMA | http://10.1001/jamainternmed.2020.4130 | Descriptive | Ecologic | Field data collection | Seroprevelance of SARS-CoV-2 antibodies by time and region | General community | General population | | | | × | |
| July | Razzaghi | Estimated County-Level Prevalence of Selected Underlying Medical Conditions Associated with Increased Risk for Severe COVID-19 Illness — United States, 2018 | MMWR | http://dx.doi.org/10.15585/mmwr.mm6929a1 | Analytic | Ecologic | Survey | Estimates of prevalence of selected underlying medical conditions | General community | General population | | | | | |
| July | Biggs | Estimated Community Seroprevalence of SARS-CoV-2 Antibodies — Two Georgia Counties, April 28–May 3, 2020 | MMWR | http://dx.doi.org/10.15585/mmwr.mm6929e2external icon | Descriptive | Ecologic | Active surveillance program | Prevelence of antibody positivity by demographic characterisitic | General community | General population | | | | × | |
| July | Menachemi | Population Point Prevalence of SARS-CoV-2 Infection Based on a Statewide Random Sample — Indiana, April 25–29, 2020 | MMWR | http://dx.doi.org/10.15585/mmwr.mm6929e1 | Descriptive | Cross-sectional | Active surveillance program | Prevelence of antibody positivity by current infection status | General community | General population | | | | × | |

Table S3. List of reviewed CDC-affiliated studies.

| | | | | | | | | | | | | | | | | | |
|---|---|---|---|---|---|---|---|---|---|---|---|---|---|---|---|---|---|
| July | Yousaf | A Prospective Cohort Study in Nonhospitalized Household Contacts With Severe Acute Respiratory Syndrome Coronavirus 2 Infection: Symptom Profiles and Symptom Change Over Time | CID | https://doi.org/10.1093/cid/ciaa1072 | Analytic | Prospective cohort | Field data collection | Prevalence of symtpoms in exposed contacts with postive RT-PCR tests | General community | General population | | | | × | | | |
| July | Tenforde | Symptom Duration and Risk Factors for Delayed Return to Usual Health Among Outpatients with COVID-19 in a Multistate Health Care Systems Network — United States, March–June 2020 | MMWR | http://dx.doi.org/10.15585/mmwr.mm6930e1external icon | Analytic | Cross-sectional | Survey | Odds of not returning to "usual health" at 14–21 days after testing | General community | General population | | | × | | | | |
| July | Paradis | Notes from the Field: Public Health Efforts to Mitigate COVID-19 Transmission During the April 7, 2020 Election — City of Milwaukee, Wisconsin, March 13–May 5, 2020 | MMWR | http://dx.doi.org/10.15585/mmwr.mm6930a4external icon | Descriptive | Ecologic | Passive surveillance program | Trends in number of reported COVID-19 cases, hospitalizations, and associated deaths | General community | General population | | | | | | | |
| July | Langdon-Embry | Notes from the Field: Rebound in Routine Childhood Vaccine Administration Following Decline During the COVID-19 Pandemic — New York City, March 1–June 27, 2020 | MMWR | http://dx.doi.org/10.15585/mmwr.mm6930a3external icon | Descriptive | Ecologic | Passive surveillance program | Trends in routine childhood vaccinations | General community | General population | | | | | | | × |
| August | Donahue | Notes from the Field: Characteristics of Meat Processing Facility Workers with Confirmed SARS-CoV-2 Infection — Nebraska, April–May 2020 | MMWR | http://dx.doi.org/10.15585/mmwr.mm6931a3external icon | Descriptive | Cross-sectional | Field data collection | Propotions of infected indivduals with demographic, household, and clinical characteristics | Industrial facility | Industrial workers | | | | | | | |
| August | Steinberg | COVID-19 Outbreak Among Employees at a Meat Processing Facility — South Dakota, March–April 2020 | MMWR | http://dx.doi.org/10.15585/mmwr.mm6931a2 | Descriptive | Case series or cluster | Field data collection | Propotions of infected indivduals with demographic and clinical characteristics; weekly cumulative attack rates | Industrial facility | Industrial workers | | | | | | | |
| August | Szablewski | SARS-CoV-2 Transmission and Infection Among Attendees of an Overnight Camp — Georgia, June 2020 | MMWR | http://dx.doi.org/10.15585/mmwr.mm6931e1 | Descriptive | Case series or cluster | Field data collection | Attack rates by demographic characteristic | Non-school youth program | Children | | | | | | | |
| August | Krueger | Characteristics and Outcomes of Contacts of COVID-19 Patients Monitored Using an Automated Symptom Monitoring Tool — Maine, May–June 2020 | MMWR | http://dx.doi.org/10.15585/mmwr.mm6931e2external icon | Descriptive | Cross-sectional | Field data collection | Propotions of infected indivduals with demographic and clinical characteristics | General community | General population | | | | | | | |
| August | Plucinski | COVID-19 in Americans aboard the Diamond Princess cruise ship | CID | https://doi.org/10.1093/cid/ciaa1180 | Descriptive | Case series or cluster | Field data collection | | Cruise ship | General population | | | | | | | |
| August | Czeisler | Mental Health, Substance Use, and Suicidal Ideation During the COVID-19 Pandemic — United States, June 24–30, 2020 | MMWR | http://dx.doi.org/10.15585/mmwr.mm6932a1external icon | Analytic | Cross-sectional | Survey | Prevelence estimates of mental health symtpoms and conditions; prevlence ratios mental health symptoms and conditions for demographic and occupational characteristics | General community | General population | | | | | | | × |
| August | Bigelow | Transmission of SARS-CoV-2 Involving Residents Receiving Dialysis in a Nursing Home — Maryland, April 2020 | MMWR | http://dx.doi.org/10.15585/mmwr.mm6932e4external icon | Descriptive | Case series or cluster | Field data collection | Proportion of infected and uninfected individuals in each residential location and with each dialysis schedule | Skilled nursing facilities | LCTF residents | | | | | | | |
| August | Godfred-Cato | COVID-19–Associated Multisystem Inflammatory Syndrome in Children — United States, March–July 2020 | MMWR | http://dx.doi.org/10.15585/mmwr.mm6932e2external icon | Descriptive | Cross-sectional | Passive surveillance program | Proportion of infected individuals with clinical characteristics | General community | Children | | | | | | | |
| August | Sutton | Notes from the Field: Seroprevalence Estimates of SARS-CoV-2 Infection in Convenience Sample — Oregon, May 11–June 15, 2020 | MMWR | http://dx.doi.org/10.15585/mmwr.mm6932a4 | Descriptive | Cross-sectional | Field data collection | Seroprevalence estimates by age | General community | General population | | | | | | × | |
| August | Hatfield | Facility-Wide Testing for SARS-CoV-2 in Nursing Homes — Seven U.S. Jurisdictions, March–June 2020 | MMWR | http://dx.doi.org/10.15585/mmwr.mm6932e5external icon | Descriptive | Ecologic | Field data collection | Prevelence of infection by testing strategy | Skilled nursing facilities | LCTF residents | | | | | | × | |
| August | Kim | Hospitalization Rates and Characteristics of Children Aged <18 Years Hospitalized with Laboratory-Confirmed COVID-19 — COVID-NET, 14 States, March 1–July 25, 2020 | MMWR | http://dx.doi.org/10.15585/mmwr.mm6932e3 | Descriptive | Cross-sectional | Active surveillance program | Cumulative incidence of hospitalization | General community | Children | | | | | | × | |
| August | Lewis | Household Transmission of SARS-CoV-2 in the United States | CID | https://doi.org/10.1093/cid/ciaa1166 | Analytic | Prospective cohort | Field data collection | SIR; odds of infection for selected contact and cllnical characteristics | General community | General population | × | × | | | | | |
| August | Bui | Racial and Ethnic Disparities Among COVID-19 Cases in Workplace Outbreaks by Industry Sector — Utah, March 6–June 5, 2020 | MMWR | http://dx.doi.org/10.15585/mmwr.mm6933e3 | Analytic | Ecologic | Passive surveillance program | Proportion of workplace outbreak versus non-outbreak infected individuals with selected risk factors | Industrial facility | Industrial workers | | × | | | | | |
| August | Davalantes | Notes from the Field: COVID-19 Prevention Practices in State Prisons — Puerto Rico, 2020 | MMWR | http://dx.doi.org/10.15585/mmwr.mm6933a4 | Descriptive | Cross-sectional | Passive surveillance program | Enumeration of infected indiividuals | Correctional or detention facilties | Incarcerated individuals | | | | | | | |

Table S3. List of reviewed CDC-affiliated studies.

| | | | | | | | | | | | | | |
|---|---|---|---|---|---|---|---|---|---|---|---|---|---|
| August | Hagan | Mass Testing for SARS-CoV-2 in 16 Prisons and Jails — Six Jurisdictions, United States, April–May 2020 | MMWR | http://dx.doi.org/10.15585/mmwr.mm6933a3external icon | Descriptive | Ecologic | Active surveillance program | Point prevelance of infection | Correctional or detention facilities | Incarcerated individuals | | X | |
| August | Oster | Trends in Number and Distribution of COVID-19 Hotspot Counties — United States, March 8–July 15, 2020 | MMWR | http://dx.doi.org/10.15585/mmwr.mm6933e2external icon | Descriptive | Ecologic | Passive surveillance program | Number of COVID-19 hotspot counties by time period | General community | General population | | | |
| August | Moore | Disparities in Incidence of COVID-19 Among Underrepresented Racial/Ethnic Groups in Counties Identified as Hotspots During June 5–18, 2020 — 22 States, February–June 2020 | MMWR | http://dx.doi.org/10.15585/mmwr.mm6933e1external icon | Descriptive | Ecologic | Passive surveillance program | Ratio of proportion of ethnic group with infection to proportion of ethnic group in population | General community | General population | | | |
| August | Link-Gelles | Limited Secondary Transmission of SARS-CoV-2 in Child Care Programs — Rhode Island, June 1–July 31, 2020 | MMWR | http://dx.doi.org/10.15585/mmwr.mm6934e2external icon | Descriptive | Case series or cluster | Active surveillance program | Enumeration of infected indiividuals | Childcare facilities | General population | | | |
| August | Hatcher | COVID-19 Among American Indian and Alaska Native Persons — 23 States, January 31–July 3, 2020 | MMWR | http://dx.doi.org/10.15585/mmwr.mm6934e1external icon | Descriptive | Cross-sectional | Passive surveillance program | Cumulative incidence of infection by AI/NA status | General community | General population | | | |
| August | McBee | Notes from the Field: Universal Statewide Laboratory Testing for SARS-CoV-2 in Nursing Homes — West Virginia, April 21–May 8, 2020 | MMWR | http://dx.doi.org/10.15585/mmwr.mm6934a4external icon | Descriptive | Ecologic | Active surveillance program | Point prevalence of infection | Skilled nursing facilities | LCTF residents | | X | |
| September | Moreland | Timing of State and Territorial COVID-19 Stay-at-Home Orders and Changes in Population Movement — United States, March 1–May 31, 2020 | MMWR | http://dx.doi.org/10.15585/mmwr.mm6935a2external icon | Analytic | Ecologic | Other | Temporal changes in mobility measure | General community | General population | | | |
| September | Blaisdell | Preventing and Mitigating SARS-CoV-2 Transmission — Four Overnight Camps, Maine, June–August 2020 | MMWR | http://dx.doi.org/10.15585/mmwr.mm6935e1external icon | Descriptive | Ecologic | Field data collection | Prevelance of characteristics of summer camp attendees | Non-school youth program | Children | | | |
| September | Self | Seroprevalence of SARS-CoV-2 Among Frontline Health Care Personnel in a Multistate Hospital Network — 13 Academic Medical Centers, April–June 2020 | MMWR | http://dx.doi.org/10.15585/mmwr.mm6935e2 | Descriptive | Cross-sectional | Field data collection | Seroprevalence | Healthcare facility | Healthcare workers | | X | |
| September | Yoon | Prevalence among People Experiencing Homelessness and Homelessness Service Staff during Early Community Transmission in Atlanta, Georgia, April–May 2020 | CID | https://doi.org/10.1093/cid/ciaa1340 | Analytic | Cross-sectional | Field data collection | Point prevalence of SARS-CoV-2 RT-PCR positive test by "sheltered" status | Homeless shelter | Homeless individuals | X | | |
| September | Fisher | Community and Close Contact Exposures Associated with COVID-19 Among Symptomatic Adults ≥18 Years in 11 Outpatient Health Care Facilities — United States, July 2020 | MMWR | http://dx.doi.org/10.15585/mmwr.mm6936a5 | Analytic | Case-control | Survey | Odds ratios for risk factors for infection | General community | General population | X | | |
| September | Silver | Prevalence of Underlying Medical Conditions Among Selected Essential Critical Infrastructure Workers — Behavioral Risk Factor Surveillance System, 31 States, 2017–2018 | MMWR | http://dx.doi.org/10.15585/mmwr.mm6936a3external icon | Analytic | Cross-sectional | Survey | Adjusted prevalence ratio of underlying health conditions among essential workers | General community | General population | | | |
| September | Czeisler | Delay or Avoidance of Medical Care Because of COVID-19–Related Concerns — United States, June 2020 | MMWR | http://dx.doi.org/10.15585/mmwr.mm6936a4 | Analytic | Cross-sectional | Survey | Adjusted prevalence ratios for characteristics associated with delay or avoidance of urgent or emergency medical care | General community | General population | | | X |
| September | Telford | Preventing COVID-19 Outbreaks in Long-term Care Facilities Through Preemptive Testing of Residents and Staff Members — Fulton County, Georgia, March–May 2020 | MMWR | http://dx.doi.org/10.15585/mmwr.mm6937a4external icon | Analytic | Ecologic | Field data collection | Prevelence of infection indentified by testing by testing strategy | Skilled nursing facilities | LCTF residents | X | | |
| September | Bui | Association Between CMS Quality Ratings and COVID-19 Outbreaks in Nursing Homes — West Virginia, March 17–June 11, 2020 | MMWR | http://dx.doi.org/10.15585/mmwr.mm6937a5external icon | Analytic | Ecologic | Passive surveillance program | Odds of skilled nursing facility outbreak by facility characteristic | Skilled nursing facilities | LCTF residents | X | | |
| September | Taylor | Serial Testing for SARS-CoV-2 and Virus Whole Genome Sequencing Inform Infection Risk at Two Skilled Nursing Facilities with COVID-19 Outbreaks — Minnesota, April–June 2020 | MMWR | http://dx.doi.org/10.15585/mmwr.mm6937a3 | Descriptive | Case series or cluster | Field data collection | Characteristics of infected individuals | Skilled nursing facilities | LCTF residents | | | |
| September | Lopez | Transmission Dynamics of COVID-19 Outbreaks Associated with Child Care Facilities — Salt Lake City, Utah, April–July 2020 | MMWR | http://dx.doi.org/10.15585/mmwr.mm6937e3external icon | Descriptive | Case series or cluster | Field data collection | Frequency of characteristics of all staff members, attendees, and their contacts associated with COVID-19 outbreaks at three child care facilities | Childcare facilities | General population | | | |
| September | Bixler | SARS-CoV-2–Associated Deaths Among Persons Aged <21 Years — United States, February 12–July 31, 2020 | MMWR | http://dx.doi.org/10.15585/mmwr.mm6937e4external icon | Descriptive | Cross-sectional | Passive surveillance program | Frequency of demographic and clinical characteristics of SARS-CoV-2–associated deaths among persons aged <21 years | General community | Children | | X | |

Table S3. List of reviewed CDC-affiliated studies.

| | | | | | | | | | | | | |
|---|---|---|---|---|---|---|---|---|---|---|---|---|
| September | Jackson | Predictors at admission of mechanical ventilation and death in an observational cohort of adults hospitalized with COVID-19 | CID | https://doi.org/10.1093/cid/ciaa1459 | Analytic | Retrospective cohort | Medical records | Odds of death for demographic and clinical predictors | General community | General population | | × |
| September | Lewis | Disparities in COVID-19 Incidence, Hospitalizations, and Testing, by Area-Level Deprivation — Utah, March 3–July 9, 2020 | MMWR | http://dx.doi.org/10.15585/mmwr.mm6938a4 | Analytic | Ecologic | Passive surveillance program | Confirmed case and hospital admissions incidence by area level deprivation measure | General community | General population | × | × |
| September | Hughes | Update: Characteristics of Health Care Personnel with COVID-19 — United States, February 12–July 16, 2020 | MMWR | http://dx.doi.org/10.15585/mmwr.mm6938a3external icon | Descriptive | Cross-sectional | Passive surveillance program | Frequency of demographic and clinical characteristics, hospitalization and intensive care unit (ICU) status among health care personnel (HCP) with confirmed COVID-19 infection by vital status | General community | Healthcare workers | | |
| September | Lash | COVID-19 Contact Tracing in Two Counties — North Carolina, June–July 2020 | MMWR | http://dx.doi.org/10.15585/mmwr.mm6938e3 | Descriptive | Ecologic | Active surveillance program | Description of contact tracing metrics in two counties | General community | General population | | |
| September | anagiotakopoulc | SARS-CoV-2 Infection Among Hospitalized Pregnant Women: Reasons for Admission and Pregnancy Characteristics — Eight U.S. Health Care Centers, March 1–May 30, 2020 | MMWR | http://dx.doi.org/10.15585/mmwr.mm6938e2external icon | Descriptive | Cross-sectional | Passive surveillance program | Frequency of demographic, illness, and pregnancy characteristics of pregnant women hospitalized with SARS-CoV-2 infection | General community | Pregnant women | | |
| September | Delahoy | Characteristics and Maternal and Birth Outcomes of Hospitalized Pregnant Women with Laboratory-Confirmed COVID-19 — COVID-NET, 13 States, March 1–August 22, 2020 | MMWR | http://dx.doi.org/10.15585/mmwr.mm6938e1 | Descriptive | Cross-sectional | Passive surveillance program | Frequency of characteristics and birth outcomes of hospitalized pregnant women with COVID-19 infectio | General community | Pregnant women | | |
| October | Wilson | Multiple COVID-19 Clusters on a University Campus — North Carolina, August 2020 | MMWR | http://dx.doi.org/10.15585/mmwr.mm6939e3 | Descriptive | Case series or cluster | Passive surveillance program | Numbers of confirmed COVID-19 cases among university students, faculty, and staff members | College or university | General population | | |
| October | Salvatore | Recent Increase in COVID-19 Cases Reported Among Adults Aged 18–22 Years — United States, May 31–September 5, 2020 | MMWR | http://dx.doi.org/10.15585/mmwr.mm6939e4 | Descriptive | Cross-sectional | Passive surveillance program | Temporal changes in weekly COVID-19 RT-PCR testing volume and confirmed case incidence by age group | General community | General population | | |
| October | Leeb | COVID-19 Trends Among School-Aged Children — United States, March 1–September 19, 2020 | MMWR | http://dx.doi.org/10.15585/mmwr.mm6939e2external icon | Descriptive | Cross-sectional | Passive surveillance program | Frequency of demographic and clinical characteristics of school-aged children aged 5–11 years and 12–17 years with positive test results for SARS-CoV-2; weekly confirmed case incidence | General community | Children | | |
| October | Boehmer | Changin+[@Publication]g Age Distribution of the COVID-19 Pandemic — United States, May–August 2020 | MMWR | http://dx.doi.org/10.15585/mmwr.mm6939e1external icon | Descriptive | Cross-sectional | Passive surveillance program | Confirmed case incidence by age and month; trends in median age of individuals with tests, postive tests, confirmed cases, and ED visits for coronavirus like illness | General community | General population | | |
| October | Haston | Characteristics Associated with Adults Remembering to Wash Hands in Multiple Situations Before and During the COVID-19 Pandemic — United States, October 2019 and June 2020 | MMWR | http://dx.doi.org/10.15585/mmwr.mm6940a2 | Analytic | Cross-sectional | Survey | Prevalence of handwashing behavior by demographic and geographical characteristic | General community | General population | | |
| October | Morris | Case Series of Multisystem Inflammatory Syndrome in Adults Associated with SARS-CoV-2 Infection — United Kingdom and United States, March–August 2020 | MMWR | http://dx.doi.org/10.15585/mmwr.mm6940e1 | Descriptive | Cross-sectional | Passive surveillance program | Summary of demographics, clinical features, treatments, and outcomes of nine adults reported to CDC with multisystem inflammatory syndrome (MIS) associated with SARS-CoV-2 infection | General community | General population | | |
| October | Schwartz | Adolescent with COVID-19 as the Source of an Outbreak at a 3-Week Family Gathering — Four States, June–July 2020 | MMWR | http://dx.doi.org/10.15585/mmwr.mm6940e2 | Descriptive | Case series or cluster | Field data collection | Number of confirmed, probable, and suspected COVID-19 cases among event participants | Private social gathering | General population | | |
| October | Galloway | Trends in COVID-19 Incidence After Implementation of Mitigation Measures — Arizona, January 22–August 7, 2020 | MMWR | http://dx.doi.org/10.15585/mmwr.mm6940e3external icon | Descriptive | Ecologic | Passive surveillance program | Timing of community mitigation measures relative to COVID-19 confirmed case counts | General community | General population | | |
| October | Czeisler | Demographic Characteristics, Experiences, and Beliefs Associated with Hand Hygiene Among Adults During the COVID-19 Pandemic — United States, June 24–30, 2020 | MMWR | http://dx.doi.org/10.15585/mmwr.mm6941a3external icon | Analytic | Cross-sectional | Survey | Prevalence of frequent hand hygiene after contact with high-touch public surfaces; odds ratios for washing hands after contact with high-touch public surfaces by respondent characteristics | General community | General population | | |
| October | Wilson | Factors Influencing Risk for COVID-19 Exposure Among Young Adults Aged 18–23 Years — Winnebago County, Wisconsin, March–July 2020 | MMWR | http://dx.doi.org/10.15585/mmwr.mm6941a3external icon | Descriptive | Cross-sectional | Passive surveillance program | Frequency of characteristics of confirmed COVID-19 cases among persons aged 18–23 years | General community | General population | | |
| October | Atrubin | An Outbreak of COVID-19 Associated with a Recreational Hockey Game — Florida, June 2020 | MMWR | http://dx.doi.org/10.15585/mmwr.mm6941a4 | Descriptive | Case series or cluster | Field data collection | Number and timeing confirmed cases among event attendees | Private social gathering | General population | | |

Table S3. List of reviewed CDC-affiliated studies.

| | | | | | | | | | | | | | | | | | |
|---|---|---|---|---|---|---|---|---|---|---|---|---|---|---|---|---|---|
| October | Oster | Transmission Dynamics by Age Group in COVID-19 Hotspot Counties — United States, April–September 2020 | MMWR | http://dx.doi.org/10.15585/mmwr.mm6941e1external icon | Descriptive | Ecologic | Passive surveillance program | Trends in the percentage of positive SARS-CoV-2RT-PCR tests by age group and U.S. Census region | General community | General population | | | | | | | |
| October | Gold | Race, Ethnicity, and Age Trends in Persons Who Died from COVID-19 — United States, May–August 2020. | MMWR | http://dx.doi.org/10.15585/mmwr.mm6942 | Descriptive | Cross-sectional | National Vital Statistics System | Propotion of deaths attributed to COVID-19 by U.S. Census region,, agegroup, and nursing home or long-term care facility status | General community | General population | | | | | | | |
| October | Cates | Risk for In-Hospital Complications Associated with COVID-19 and Influenza — Veterans Health Administration, United States, October 1, 2018–May 31, 2020. | MMWR | http://dx.doi.org/10.15585/mmwr.mm6942 | Analytic | Retrospective cohort | Medical records | Proportions and adjusted relative risk of selected COVID-19 respiratory and nonrespiratory complications for patients hospitalized with COVID vs Influenza | General community | General population | | | | | | | |
| October | Rossen | Excess Deaths Associated with COVID-19, by Age and Race and Ethnicity — United States, January 26–October 3, 2020. | MMWR | http://dx.doi.org/10.15585/mmwr.mm6942e2 | Descriptive | Cross-sectional | National Vital Statistics System | Excess deaths associated with COVID-19 status, by week, by agegroup, by race / ethnicity designation | General community | General population | | | | | | × | |
| October | Murray | Mitigating a COVID-19 Outbreak Among Major League Baseball Players — United States, 2020. | MMWR | http://dx.doi.org/10.15585/mmwr.mm6942a4 | Descriptive | Case series or cluster | Field data collection | ? | General community | Athletes | | | | | | | |
| October | Dasgupta | Association Between Social Vulnerability and a County's Risk for Becoming a COVID-19 Hotspot — United States, June 1–July 25, 2020. | MMWR | http://dx.doi.org/10.15585/mmwr.mm6942a3 | Analytic | Ecologic | Passive surveillance program | Associations between social vulnerability measures* and hotspot identification | General community | General population | | × | | | | | |
| October | Pringle | COVID-19 in a Correctional Facility Employee Following Multiple Brief Exposures to Persons with COVID-19 — Vermont, July–August 2020 | MMWR | http://dx.doi.org/10.15585/mmwr.mm6943e1external icon. | Descriptive | Case series or cluster | Field data collection | Summary of type, frequency, and duration of close contacts | Correctional or detention facilties | Correctional facility workers | | | | | | | |
| October | Kambhampati | COVID-19–Associated Hospitalizations Among Health Care Personnel — COVID-NET, 13 States, March 1–May 31, 2020. | MMWR | http://dx.doi.org/10.15585/mmwr.mm6943e3 | Descriptive | Cross-sectional | Active surveillance program | Demographic and clinical characteristics of health care personnel (HCP) with COVID-19-associated hospitalizations | Healthcare facility | Healthcare workers | | | | | | | |
| October | Teran | Outbreak Among a University's Men's and Women's Soccer Teams — Chicago, Illinois, July–August 2020. | MMWR | http://dx.doi.org/10.15585/mmwr.mm6943e5 | Analytic | Case-control | Field data collection | Odds ratios for risk factors for infection | College or university | Athletes | | × | | | | | |
| October | Hutchins | COVID-19 Mitigation Behaviors by Age Group — United States, April–June 2020 | MMWR | http://dx.doi.org/10.15585/mmwr.mm6943e4 | Analytic | Cross-sectional | Survey | Prevalence of engagement in mitigation behaviors | General community | General population | | | | | | | |
| October | Fell | SARS-CoV-2 Exposure and Infection Among Health Care Personnel — Minnesota, March 6–July 11, 2020 | MMWR | http://dx.doi.org/10.15585/mmwr.mm6943a5 | Analytic | Cross-sectional | Field data collection | Prevalence of personal protective equipment (PPE) use by health care personnel (HCP) during SARS-CoV-2 exposures | Healthcare facility | healthcare workers | | | | | | | |
| October | Pray | COVID-19 Outbreak at an Overnight Summer School Retreat — Wisconsin, July–August 2020 | MMWR | http://dx.doi.org/10.15585/mmwr.mm6943a4 | Descriptive | Case series or cluster | Field data collection | Frequency of characteristics of confirmed COVID-19 cases among event attendees | Public social gathering | Children | | | | | | | |
| October | Koonin | Trends in the Use of Telehealth During the Emergence of the COVID-19 Pandemic — United States, January–March 2020 | MMWR | http://dx.doi.org/10.15585/mmwr.mm6943a3 | Descriptive | Ecologic | Other | Trends in telehealth visits | General community | General population | | | | | | | × |
| October | Grijalva | Transmission of SARS-COV-2 Infections in Households — Tennessee and Wisconsin, April–September 2020 | MMWR | http://dx.doi.org/10.15585/mmwr.mm6944e1 | Analytic | Prospective cohort | Field data collection | SIR; by age | General community | General population | × | | | | | | |
